# Hospital strain and Covid-19 fatality: analysis of English nationwide surveillance data

**DOI:** 10.1101/2022.09.27.22280401

**Authors:** Teng-Fei Lin, Zi-Yi Zhao, Zhi-Rong Yang, Bing-li Li, Chang Wei, Fu-Xiao Li, Yi-Wen Jiang, Di Liu, Zu-Yao Yang, Feng Sha, Jin-Ling Tang

## Abstract

**Objectives:** To examine whether and to what extent hospital strain will increase the risk of death from Covid-19.

**Design:** Retrospective cohort study.

**Setting:** England.

**Participants:** Data on all the 147,276 Covid-19 deaths and 601,084 hospitalized Covid-19 patients in England during the period between 9 April 2020 and 11 March 2022 were extracted on a daily basis from the UK Health Security Agency.

**Main outcome measures:** The number of Covid-19 patients currently in hospitals was used as the measure of hospital strain. Daily case fatality was estimated as the measure of risk of death from Covid-19. The study was divided into 4 periods, which represented largely the wild, Alpha, Delta and Omicron waves. Weighted linear regression models were used to assess the association between hospital strain and Covid-19 fatality with adjustment for potential confounders including vaccination score, hospital admission rate, percentage of deaths outside hospitals, study period and interaction between patients currently in hospitals and study period.

**Results:** The daily case fatality from Covid-19 increased linearly as the number of patients currently in hospitals increased in the 4 study periods except the Omicron wave. After adjusting for potential confounders, an increase in 1000 patients currently in hospitals was associated with a relative increase of 6.3% (95% CI: 5.9%~6.8%), 1.4% (95% CI: 1.3% ~ 1.5%) and 12.7% (95% CI: 10.8%~14.7%) in daily case fatality during study periods 1, 2 and 3 respectively. Compared with the lowest number of patients currently in hospitals, the highest number was associated with a relative increase of 188.0% (95% CI: 165.9%~211.6%), 69.9% (95% CI: 59.0%~81.8%) and 58.2% (95% CI: 35.4%~89.0%) in daily case fatality in the first 3 study periods respectively. Sensitivity analyses using the number of patients in ventilation beds as the measure of hospital strain showed similar results.

**Conclusions:** The risk of death from Covid-19 was linearly associated with the number of patients currently in hospitals, suggesting any (additional) effort to ease hospital strain or maintain care quality be beneficial during large outbreaks of Covid-19 and likely of other similar infectious diseases.

**Summary box:** *What is already known on this topic:*

- During the Covid-19 pandemic, tremendous efforts have been made in many countries to suppress epidemic peaks and strengthen hospital services so as to avoid hospital strain with an ultimate aim to reduce the risk of death from Covid-19.
- These efforts were made according to the widely held belief that hospital strain would increase the risk of Covid-19 death but good empirical evidence was largely lacking to support the hypothesis.
- A few small studies showed that shortage in intensive care was associated with an increased Covid-19 fatality but strains may occur in many areas in the healthcare system besides intensive care and they may all increase the risk of death from Covid-19.
- The totality of hospital strain can be approximated by the number of patients currently in hospitals but its effects on the risk of Covid-19 death has not been demonstrated.

*What this study adds:*

- We found the risk of death from Covid-19 was linearly associated with the number of patients currently in hospitals before the Omicron period.
- Compared with the lowest number of patients currently in hospitals in an outbreak, the highest number could be associated with a relative increase in the risk of death between 58.2% and 188.0%.
- The number of patients currently in hospitals during the Omicron period was not found associated with the risk of death but there remains uncertainty if the number of patients currently in hospitals reached a level much higher than that actually occurred in England or in places other than England.

*How this study might affect research, practice, or policy:*

- Facing the on-going Covid-19 pandemic and future outbreaks alike, the linear relation between hospital strain and fatality suggests importantly any (additional) effort to reduce hospital strain would be beneficial during a large Covid-19 outbreak.

## Background

Coronavirus disease 2019 (Covid-19) has caused a once-in-a-century pandemic. As of 5 September 2022, the pandemic had resulted in 600,366,479 confirmed cases and 6,460,493 deaths worldwide ^1^. Facing the pandemic, countries adopted however polarized prevention and control policies. A so-called herd immunity policy was used in the United Kingdom and many other countries ^2,3^, whereas a dynamic clearance or zero-Covid-19 containment strategy was adopted in China mainland ^4^. Under the herd immunity policy, the epidemic was allowed to exist but in a controlled manner so that the number of cases was suppressed through public health measures to a level that could be effectively managed by available clinical resources, including in particular hospital and intensive care beds, with a belief that hospital strain can increase the risk of death from Covid-19^5,6^.

Although tremendous efforts have been made worldwide and will continue to be made to prevent hospital strain during an attack of Covid-19 outbreaks, there is limited empirical evidence that hospital strain indeed increases the risk of death from Covid-19. A few small studies reported that the hospital bed occupancy rate and hospital admission volume might be associated with the risk of death from Covid-19 ^7-9^, but their findings have yet to confirmed in large studies. Besides, a few other studies showed that strain in intensive care was associated with increased risk of death from Covid-19 ^10-14^. However, a sudden surge of a large number of cases during a Covid-19 outbreak could overwhelm not only intensive care but the entire healthcare system and cause complexities and chaos in coordination of services as well as shortages in ambulance, hospital beds, intensive care facilities, drugs, devices, testing and examination equipment and staff. These healthcare system strain may all add up and increase the risk of death but can be collectively approximated by the number of patients currently in hospitals (PIH). Thus, we used the number of PIH as the measure of hospital strain to examine its effects on the risk of death from Covid-19 in this large cohort study.

## Methods

### Study design and data sources

This is a retrospective cohort study by using data extracted on a daily basis from the UK Health Security Agency ^15^, the most comprehensive and authoritative source of data on Covid-19 in the country. The baseline data included the daily number of newly diagnosed Covid-19 cases, PIH, patients currently in ventilation beds, and cumulative vaccination rates of 1, 2 and 3 doses in people aged 12 or above. Diagnosis of cases was confirmed by nucleic acid testing and the date of reporting was that of sampling for the testing. The outcome of interest was those who died with Covid-19 on the same day.

### Definitions

#### Fatality

Daily case fatality was defined as the ratio of daily number of deaths from Covid-19 over daily number of Covid-19 PIH ^16,17^. The problem of this definition is that the number of deaths included death events that occurred outside hospitals. This issue was addressed by examining the correlation between the percentage of deaths outside hospitals and the number of PIH, and considering the percentage of deaths outside hospitals as a potential confounder in the multiple regression analyses assessing the hospital strain-fatality relation (see the statistical analysis section).

#### Hospital strain

Hospital strain was measured by the total number of PIH in a day.

#### Study periods

As the variant of the virus, vaccination coverage, and hospital strain differed considerably over time, we divided the study into four different periods and examined the effect of hospital strain on fatality separately. The first three periods were defined and divided according to the lowest numbers of cases between two major epidemic periods in England, while period 4 started from the reporting of the first Omicron case in the UK. The epidemic period before 9 April 2020 was excluded from the analyses as there was no sufficient nucleic acid testing capacity for diagnosing all infections during the period ^18^ and fatality estimates were highly likely biased.

#### Vaccination score

The vaccination rates of 1, 2 and 3 doses were converted into a single vaccination score as a measure of overall protection effect by vaccinations. Let P_1_= percentage of people having received 1 dose of vaccine, P_2_ for 2 doses, and P_3_ for 3 doses. Then the vaccination score = (P_1_−P_2_) + 2(P_2_−P_3_) + 3P_3,_ which assumes that 1 dose and 2 doses give a protection approximately 1/3 and 2/3 of that of 3 doses according to current available evidence on the protection rate of vaccination ^19^.

### Statistical analysis

The 7-day moving average of daily number of new cases, PIH and deaths, vaccination rate, and daily case fatality were described chronologically in a line chart. Summary results of these variables for the 4 periods were described in a table.

The relation of hospital strain and fatality was examined graphically by scatter-plots and by log-linear multivariate regression analyses weighted according to the daily number of PIH. Potential confounders adjusted included the vaccination score, admission rate (as an approximate measure of the severity of illness of patients upon hospital admission), the percentage of deaths outside hospitals (as an approximate quantification of the size of error in the number of deaths for estimating the fatalities), study period (as an approximate measure of the total effect of the variants of the virus, improvements in hospital care for Covid-19 patients and other unmeasured potential confounders that differed or changed over the 4 study periods), and interaction between patients in hospitals and study period. Data on the percentage of deaths outside hospitals were available on a weekly basis and acquired from the Office for National Statistics website ^20^. Besides, we are aware that PIH acts as both the independent variable and the denominator for estimating the dependent variable (the fatality). A spurious negative association can in theory arise between them. As a result, a positive PIH-fatality association will be underestimated and stronger than that thus observed.

The daily case fatality was also compared for the time points when hospitals were least and most strained (namely at the lowest and highest number of PIH) by using the simple regression of fatality against the number of PIH. The relative increase in fatality from the least to most strained time point was estimated and used to reflect the actual maximum increase in fatality due to hospital strain during a study period.

All analyses were conducted separately for the four study periods. P value ≤0.05 was considered statistically significant for all significance tests and 95% confidence intervals were constructed for all estimates. All statistical analyses were performed by using Stata software (version 17). The epidemic curves were drawn with OriginPro software (version 9.9.0.225). The scatter plots were performed in R software (version 3.6.3) by using the ggplot2 package.

### Patient and public involvement

Patients or the public were not involved in the design, or conduct, or reporting, or dissemination plans of our research.

## Results

A total of 16,498,319 confirmed Covid-19 cases, 601,084 hospitalized cases and 147,276 related deaths were reported during the 702 days of the entire study period between 9 April 2020 and 11 March 2022 in England and all included in the study. The duration of the study period, the total and median daily number of new cases and deaths, median percentage of population vaccinated with 1, 2 and 3 doses, and median daily case fatality were presented according to the 4 periods of the epidemic in Table 1.

**Table 1.** **The total number of new cases and death events, average of daily new cases, death cases and cases in hospitals, and percentage of first, second and third dose of vaccination, and daily case fatality (i.e., daily number of deaths over daily number of cases in hospital) during the 4 periods of the COVID epidemic in England between 9 April 2020 and 11 March 2022**

| Variables | Four periods of the epidemic |  |  |  | Total<br>(90/04/2020-11/03/2022) |
| --- | --- | --- | --- | --- | --- |
|  | Period 1<br>(09/04/2020-11/07/2020) | Period 2<br>(12/07/2020-30/04/2021) | Period 3<br>(01/05/2021-26/11/2021) | Period 4<br>(27/11/2021-11/03/2022) |  |
| **Daily new cases and deaths** |  |  |  |  |  |
| No. of days in the period | 94 | 293 | 210 | 105 | 702 |
| Total no. (%) of new cases | 186 230 (1.1) | 3 696 656 (22.4) | 4 855 376 (29.4) | 7 760 057 (47.0) | 16 498 319 (100.0) |
| Total no. (%) of deaths | 36 335 (24.7) | 83 400 (56.6) | 14 554 (9.9) | 12 987 (8.8) | 147 276 (100.0) |
| Total no. (%) of hospital admission | 68 382 (11.4) | 287 440 (47.8) | 111 316 (18.5) | 133 946 (22.3) | 601 084 (100.0) |
| Median (quartiles) no. of new cases | 1 425 [760, 3 125] | 7 780 [2 224, 18 197] | 25 598 [13 505, 31 601] | 62 303 [39 223, 93 784] | 15 156 [2 752, 31 327] |
| Median (quartiles) no. of deaths | 240 [91, 583] | 124 [28, 413] | 78 [18, 106] | 111 [94, 156] | 102 [39, 224] |
| Median (quartiles) of the percentage of deaths outside hospitals (%) * | 43.4 [41.2, 46.6] | 29.5 [25.2, 34.2] | 20.5 [17.9, 24.3] | 30.2 [21.8, 32.3] | 28.9 [21.7, 35.1] |
| Median (quartiles) no. of hospital admission | 562 [300, 1 058] | 528 [135, 1 467] | 636 [196, 751] | 1 220 [926, 1 604] | 696 [218, 1 232] |
| Median (quartiles) no. of patients in hospital | 7 360 [3717, 12 265] | 5 976 [1 378, 14 411] | 5 032 [1 274, 6 066] | 9 369 [7 114, 13 331] | 5 996 [2 117, 11 585] |
| Median (quartiles) no. of patients in ventilation beds | 764 [299, 1 832] | 682 [178, 1 339] | 683 [228, 810] | 608 [355, 770] | 674 [247, 910] |
| **% of population vaccinated** |  |  |  |  |  |
| Median (quartiles) completed 1<sup>st</sup> dose | 0.0 [0.0, 0.0] | 0.0 [0.0, 27.7] | 82.0 [75.3, 85.0] | 90.6 [89.7, 91.3] | 52.0 [0.0, 84.2] |
| Median (quartiles) completed 2<sup>nd</sup> dose | 0.0 [0.0, 0.0] | 0.0 [0.0, 1.0] | 69.8 [55.4, 78.0] | 83.3 [82.0, 84.6] | 4.8 [0.0, 76.8] |
| Median (quartiles) completed 3<sup>rd</sup> dose | 0.0 [0.0, 0.0] | 0.0 [0.0, 0.0] | 0.0 [0.0, 3.2] | 63.3 [56.1, 65.2] | 0.0 [0.0, 0.0] |
| **Median (quartiles) of daily case fatality (%)** | 3.4 [2.6, 4.9] | 2.7 [2.1, 3.0] | 1.6 [1.4, 1.8] | 1.2 [1.0, 1.4] | 1.9 [1.5, 2.8] |

\* Data was available on a weekly basis.

Notably, the median daily number of new cases increased steadily from 1,425 cases per day in period 1 to 62,303 cases per day in period 4 (mostly Omicron), a 43.7-folds of increase, but the daily number of PIH did not vary proportionately to the daily new cases diagnosed and showed a maximum of only 1.9-folds difference in the 4 periods. The median daily number of deaths was, however, the highest during period 1, resulting in a declining daily case fatality during the 4 periods from the highest 3.4% in period 1 to the lowest 1.2% in period 4, a 64.7% decrease (P=0.0137). The decline in fatality could only partially be explained by vaccination as there was no or only a few people who completed 2 doses of vaccination during the first 3 study periods.

The 7-day moving average of daily number of new cases, PIH and deaths, and daily case fatality during the 4 periods of the epidemic in relation to the progress of vaccination and changes in lockdowns, public health measures and variants of the virus was shown graphically in Figure 1. Patterns similar to those described above can be visually observed among daily new cases, PIH, deaths and fatality.

**Figure 1.**
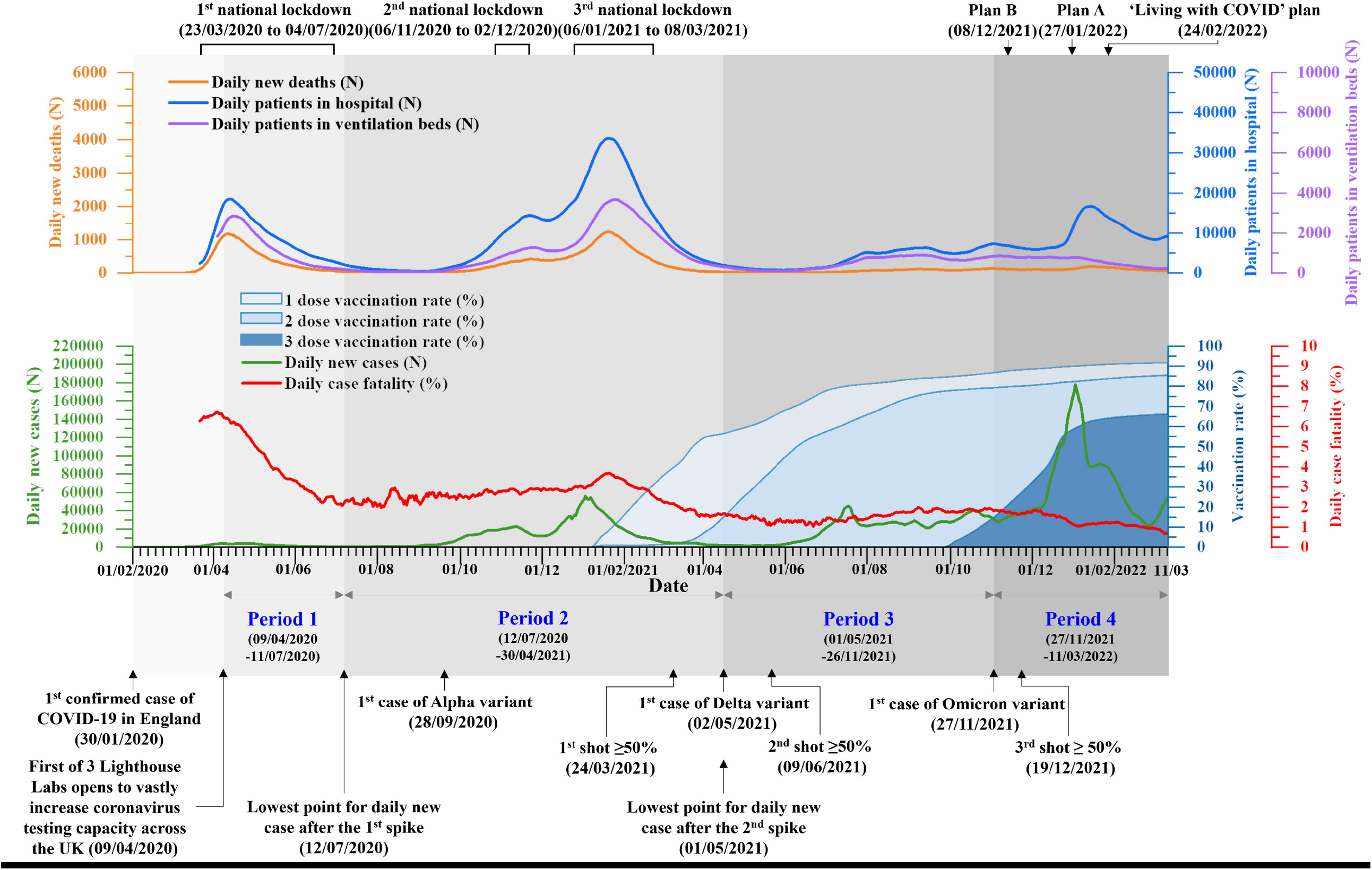
**Daily number of COVID-19 new cases, cases in hospital and death events, and daily case fatality (i.e., daily number of deaths over daily number of cases in hospital) during the 4 periods of the COVID-19 epidemic and in relation to the progress of vaccination in England between 9 April 2020 and 11 March 2022**

Importantly, the association between the daily case fatality and number of PIH, the measure of hospital strain in this study, according to the 4 periods of epidemic was shown in Figurer 2. In periods 1, 2 and 3, fatality was positively and linearly associated with the number of PIH with a correlation coefficient being 0.95, 0.55 and 0.58 respectively (all P values≤0.0001). In period 4, the fatality was sharply divided into two parts. The first part was mostly Delta and the second part was predominantly Omicron in which the fatality was the lowest and remained stable regardless of the variations in the number of PIH. The same conclusions can be drawn when patients currently in ventilation beds were used as the measure hospital strain (Appendix Figure 1). After adjusting for vaccination score, admission rate, percentage of deaths outside hospitals, study period, and interaction term between PIH and study period, hospital strain remained statistically significantly associated with daily case fatality (all P values ≤ 0.0001 in study periods 1, 2 and 3 respectively) (Table 2).

**Table 2.** **Relative increase in daily case fatality for a 1000-increase in daily number of patients in hospitals according to epidemic period and adjusted for potential confounders**

| **Study periods** | **Regression coefficients**<br>**(95% CI)\*** | **Relative increase**<br>**(95% CI)** | **P-value** |
| --- | --- | --- | --- |
| Period 1 | 0.062(0.057,0.066) | 1.063(1.059,1.068) | <0.0001 |
| Period 2 | 0.014(0.013,0.015) | 1.014(1.013,1.015) | <0.0001 |
| Period 3 | 0.120(0.102,0.137) | 1.127(1.108,1.147) | <0.0001 |
| Period 4 | -0.005(-0.014,0.003) | 0.995(0.986,1.003) | 0.2306 |

\*The model was adjusted for vaccination score, study period, interaction term between daily number of patients in hospital and study period, admission rate, and percentage of deaths outside hospitals, and weighted by number of patients in hospital (detailed results in Supplementary Table 1).

**Figure 2.**
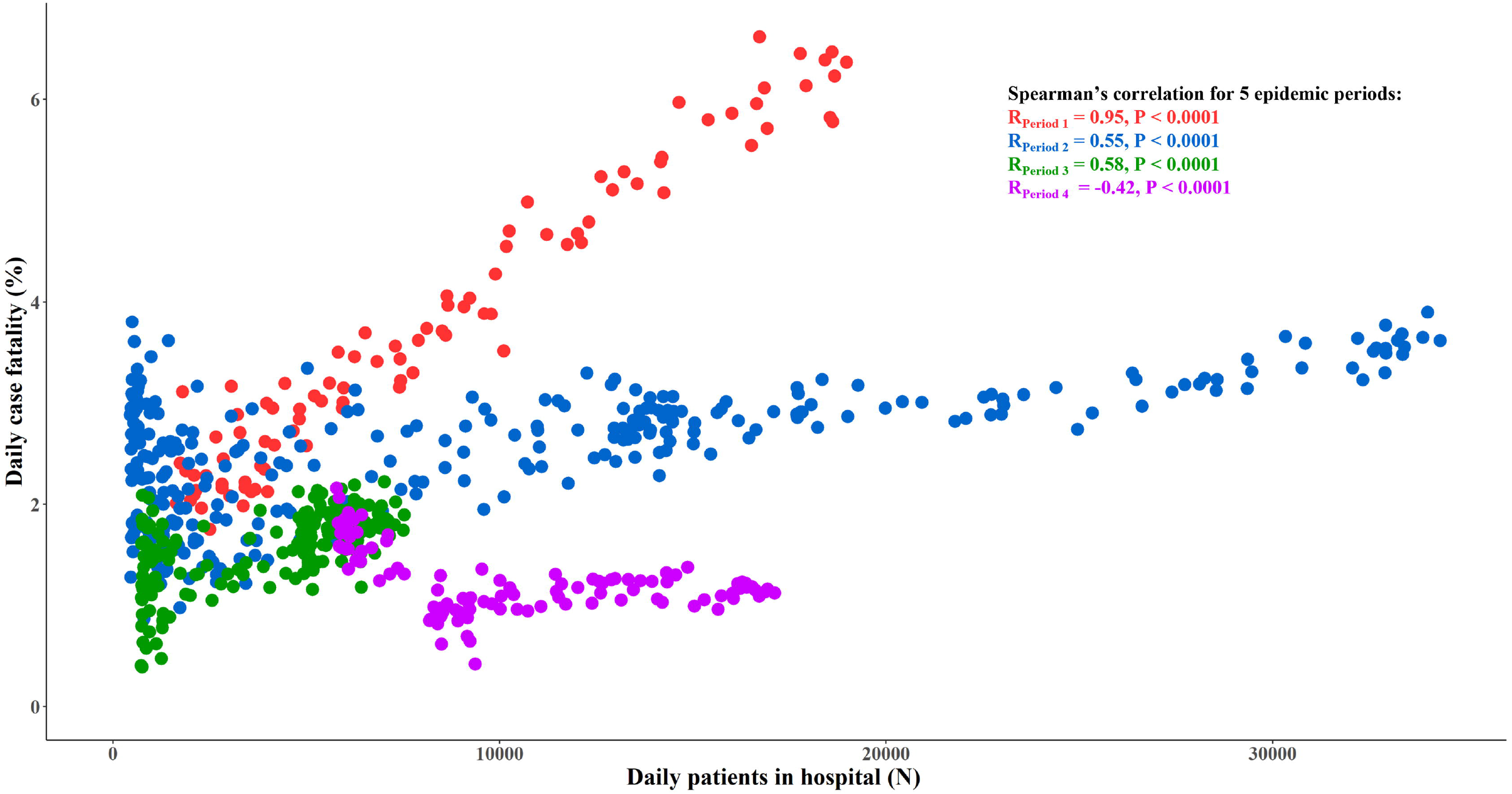
**The correlation between daily case fatality and daily number of patients in hospitals according to 4 periods of epidemic in England between 9 April 2020 and 11 March 2022**

Finally, as the daily number of PIH increased from the lowest to the highest, the actual (or unadjusted) daily case fatality was increased by 188.0% (95% CI: 165.9%~211.6%), 69.9% (95% CI: 59.0%~81.8%) and 58.2% (95% CI: 35.4%~89.0%) respectively in study periods 1, 2 and 3 (Table 3).

**Table 3.** **Percentage increase in daily case fatality when the actual daily number of patients in hospital was increased from the lowest to the highest in each of the 4 epidemic periods**

| Study period | Daily number of patients in hospitals (N) |  | Estimated daily case fatality (%) * |  |  |
| --- | --- | --- | --- | --- | --- |
|  | Lowest | Highest | At the lowest number patients in hospital (a) | At the highest number patients in hospital (b) | Percentage increase (b-a)/a # |
| Period 1 | 1640 | 18974 | 2.39(2.29,2.49) | 6.87(6.65,7.10) | 188.0(165.9,211.6) |
| Period 2 | 451 | 34336 | 2.15(2.09,2.21) | 3.65(3.56,3.74) | 69.9(59.0,81.8) |
| Period 3 | 730 | 7535 | 1.22(1.14,1.30) | 1.93(1.86,2.00) | 58.2(35.4,89.0) |
| Period 4 | 5784 | 17120 | 1.25(1.15,1.36) | 1.08(1.00,1.17) | -13.5(-24.0,0.5) |

\* Estimated by using simple linear regression of log daily case fatality against daily number of patients in hospitals, weighted by daily number of patients in hospitals.

## Discussion

By using authoritative English national data over 2 years, we found the daily number of Covid-19 PIH as an indicator of overall hospital strain the epidemic caused was linearly associated with the risk of death from Covid-19 except in the Omicron period, which confirmed findings from several previous studies^7-9^. The largest difference in the risk of death from Covid-19 observed during an outbreak in England was 2.88 folds, suggesting theoretically a maximum of 65.3% death risk reduction be achievable by reducing Covid-19 PIH but the linear relation suggests that any additional effort to reduce Covid-19 PIH is related to a reduction in the risk of death and worthwhile regardless of the total number of hospital beds available and its percentage occupied. Our findings provide strong evidence that lands support for efforts to ease hospital strain so as to reduce deaths from Covid-19 outbreaks and have important implications for the on-going Covid-19 pandemic and very likely also for any similar infectious disease outbreaks in the future.

The number of Covid-19 PIH is a composite indicator for overall hospitals strain, which can be caused by many inter-related factors within and outside hospitals in a complex manner. The hospitals factors include personnel, facilities, equipment, drugs, ventilation beds, and their preparedness. Non-pharmacological interventions (NPIs) and vaccination are the major efforts that can be mobilized outside hospitals to reduce hospital strain^21-23^. In addition, factors such as the variant of the virus and patients’ care seeking behaviors also affect hospital strain. For example, shortage in intensive care was shown associated with an increased risk of death from Covid-19 in the early stage of the pandemic in various countries ^10-14^. Our analyses with a much larger data set also showed that the number of ventilation beds Covid-19 patients occupied had a similar effect on fatality. However, studies on these individual determinants of hospital strain may underrate the effect of overall hospital strain on Covid-19 fatality as these studies are restricted only to a small fraction of all patients who may die^10,24^.

Furthermore, these factors may work together to cause difficulties for patients to be admitted to hospitals ^25^, infections in hospital staff ^26^, and inpatient cross-infections ^27^, which in turn further increases hospital strain. Importantly, most of these factors and their interactions in a given place or setting would change dynamically over time. Thus, different profiles of hospital strain determinants in a place during different periods of the epidemic may well explain the different patterns of the hospital strain-fatality relation found in our study. For example, hospitals were least prepared at the beginning of the pandemic and as a result the highest fatality was observed during period 1 in our study. As hospitals gained more experiences and became more prepared, the hospital strain-fatality relation had gradually become less evident. During the last period of our study, the Omicron variant caused least severe infections ^28,29^, the majority of people had been vaccinated ^15^, almost all patients in UK hospitals had been routinely tested for antigen and hospitals, care management and NPIs had become most prepared and efficient ^30^. Consequently, the number of PIH during this period was maintained at a relatively low level and below it hospital strain was not shown related to fatality.

If the pressure on medical facilities and staff is reduced by freeing up appropriately staffed beds, the health care for Covid-19 patients may improve and the risk of death be reduced. Polices including building temporary facilities (e.g. the Nightingale hospitals in the UK), cancelation of elective admissions, and a stricter triage of new admissions and management of mild to moderate cases in the community have been implemented to ease hospital strain. This study also shows how the relationship between hospital strain and case fatality varied with the different viral variants, providing further implications for policy-making. Having said that, we would like to emphasize that England’s experiences with Omicron may not apply to Omicron outbreaks in all other places. If NPIs were not mobilized quickly and sufficiently, outbreaks of Omicron variants can still raise the number of patients in hospitals to a level that is high enough to increase the risk of death from Covid-19 as happened in early 2022 in Hong Kong which experienced one of the highest fatality rates from Omicron outbreaks in the world ^31,32^.

Out study is based on a large amount of data from England, which collected and maintained comprehensive and high quality data on Covid-19, deaths related and other relevant factors. Although we have made tremendous efforts to reduce biases and control for confounding, the hospital strain-fatality relation may still be fully or partly explained by residual biases and confounding. We observed that the fatality was higher when the numbers of PIH was larger. Understandably, as the total number of hospital beds was relatively stable during the period of an outbreak and thus only a fixed number of patients could be admitted. It is therefore possible that more severe patients were admitted when a large number of patients needed to be admitted. Consequently, the number of PIH would be positively related to the severity of patients, causing a false relation between hospital strain and fatality. The admission rate of all Covid-19 patients can be used as an indicator for the severity of patients admitted and it indeed varied considerably, ranging from 0.8% to 63.4% during the 4 study periods. We found the admission rate was not adversely associated with the number of PIH within the study periods (Appendix Figure 2). Moreover, the hospital strain-fatality relation was affected little when admission rate was included in multiple regression analyses. Thus, we believe that the hospital strain-fatality relation is unlikely a result of severity of patients admitted.

Second, our main analyses used the daily number of deaths divided by the number of PIH as the risk of death. However, the daily numbers of PIH may not be completely comparable in their severity as they may have different amalgamations of patients at different stages of disease. It is likely that the percentage of patients admitted in earlier days and stayed on in hospitals was relatively smaller when a large number of patients need to be admitted in recent days. As patients admitted in earlier days and stayed on in hospitals were likely more severe than those admitted in recent days ^33^, the hospital strain-fatality relation in our study would have been underestimated as a result.

Third, the numerator of the daily case fatality included an average of 28.7% of deaths outside hospitals. This may lead to overestimation of the fatality at the peaks of outbreaks when a larger proportion of patients could not be admitted to hospitals and some of them died, causing a false hospital strain-fatality relation. However, the hospital strain-fatality relation was unlikely biased by the deaths outside hospitals. First, the percentage of deaths outside hospitals was not positively associated with the number of PIH, implying that it was similar regardless of the number of PIH and could not have caused a bias on the hospital strain-fatality relation (Appendix Figure 3). In addition, the result of the hospital strain-fatality relation was not changed after the percentage of deaths outside hospitals was included in multiple regression analyses.

Finally, the variant of the virus, the number of ventilation beds available and vaccination rate could all change over time and caused confounding bias in the relation between the number of PIH and fatality. However, we believed that by dividing into four study periods confounding effects by the variant of the virus and the number of ventilation beds available have been reduced to a minimum as they had either not changed or changed only slightly within a study period. The confounding effect of vaccination rate was ruled out by including it in multiple regression analyses.

## Conclusions

Hospital strain is linearly associated with the risk of death from Covid-19, suggesting any (additional) efforts to ease hospital strain be beneficial. NPIs, vaccination, and hospital preparedness should be used in joint force to minimize deaths from the on-going pandemic.

## Data Availability

All raw data included in this study were derived from publicly available documents cited in the references. Extracted and generated data are available upon request to the corresponding author.

## Author Contributions

JLT, FS, ZZY, ZYY and TFL conceived the research question and designed the study. TFL and ZZY conducted the literature search. TFL, ZYZ, BLL and CW performed the data analyses. JLT, FS, ZYY, ZRY, and TFL contributed to the interpretation of the results. TFL and ZRY wrote the first draft of the manuscript. JLT, FS, ZYY, BLL, CW, FXL, YWJ, DL critically revised the manuscript. All authors acknowledge full responsibility for the analyses and interpretation of the report. All authors have read and approved the final manuscript. FS and JLT is the guarantor. The corresponding authors attest that all listed authors meet authorship criteria and that no others meeting the criteria have been omitted.

## Reference

1. WHO Coronavirus (COVID-19) Dashboard. https://covid19.who.int/ (accessed September 2022).

2. Omer SB, Yildirim I, Forman HP. Herd Immunity and Implications for SARS-CoV-2 Control. JAMA 2020; 324(20): 2095-6.

3. Xia Y, Zhong L, Tan J, et al. How to Understand “Herd Immunity” in COVID-19 Pandemic. Front Cell Dev Biol 2020; 8: 547314.

4. Chen Q, Rodewald L, Lai S, Gao GF. Rapid and sustained containment of covid-19 is achievable and worthwhile: implications for pandemic response. BMJ 2021; 375: e066169.

5. Davies NG, Kucharski AJ, Eggo RM, Gimma A, Edmunds WJ, Centre for the Mathematical Modelling of Infectious Diseases C-wg. Effects of non-pharmaceutical interventions on COVID-19 cases, deaths, and demand for hospital services in the UK: a modelling study. Lancet Public Health 2020; 5(7): e375–e85.

6. Li R, Rivers C, Tan Q, Murray MB, Toner E, Lipsitch M. Estimated Demand for US Hospital Inpatient and Intensive Care Unit Beds for Patients With COVID-19 Based on Comparisons With Wuhan and Guangzhou, China. JAMA Netw Open 2020; 3(5): e208297.

7. Castagna F, Xue X, Saeed O, et al. Hospital bed occupancy rate is an independent risk factor for COVID-19 inpatient mortality: a pandemic epicentre cohort study. BMJ Open 2022; 12(2): e058171.

8. Karaca-Mandic P, Sen S, Georgiou A, Zhu Y, Basu A. Association of COVID-19-Related Hospital Use and Overall COVID-19 Mortality in the USA. J Gen Intern Med 2020.

9. Soria A, Galimberti S, Lapadula G, et al. The high volume of patients admitted during the SARS-CoV-2 pandemic has an independent harmful impact on in-hospital mortality from COVID-19. PLoS One 2021; 16(1): e0246170.

10. Bravata DM, Perkins AJ, Myers LJ, et al. Association of Intensive Care Unit Patient Load and Demand With Mortality Rates in US Department of Veterans Affairs Hospitals During the COVID-19 Pandemic. JAMA Netw Open 2021; 4(1): e2034266.

11. Mojoli F, Cutti S, Mongodi S, et al. The potential role of ICU capacity strain in COVID-19 mortality: comparison between first and second waves in Pavia, Italy.

12. Wilcox ME, Rowan KM, Harrison DA, Doidge JC. Does Unprecedented ICU Capacity Strain, As Experienced During the COVID-19 Pandemic, Impact Patient Outcome? Crit Care Med 2022; 50(6): e548–e56.

13. Wilde H, Mellan T, Hawryluk I, et al. The association between mechanical ventilator compatible bed occupancy and mortality risk in intensive care patients with COVID-19: a national retrospective cohort study. BMC Med 2021; 19(1): 213.

14. Guillon A, Laurent E, Duclos A, et al. Case fatality inequalities of critically ill COVID-19 patients according to patient-, hospital- and region-related factors: a French nationwide study. Ann Intensive Care 2021; 11(1): 127.

15. GOV.UK Coronavirus (COVID-19) in the UK. https://coronavirus.data.gov.uk/ (accessed March 2022).

16. Rothman K J GS, Lash T L.. Modern epidemiology, 2nd edition.: Philadelphia: Wolters Kluwer Health/Lippincott Williams & Wilkins; 1998.

17. Fletcher RHaF, S.W. and Fletcher, G.S. Clinical epidemiology: The essentials: Fifth edition; 2013.

18. Health Secretary launches biggest diagnostic lab network in British history to test for coronavirus. https://www.gov.uk/government/news/health-secretary-launches-biggest-diagnostic-lab-network-in-british-history-to-test-for-coronavirus (accessed July 2022).

19. Au WY, Cheung PP. Effectiveness of heterologous and homologous covid-19 vaccine regimens: living systematic review with network meta-analysis. BMJ 2022; 377: e069989.

20. Deaths registered weekly in England and Wales, provisional. https://www.ons.gov.uk/peoplepopulationandcommunity/birthsdeathsandmarriages/deaths/datasets/weeklyprovisionalfiguresondeathsregisteredinenglandandwales (accessed July 2022).

21. Wilder-Smith A, Freedman DO. Isolation, quarantine, social distancing and community containment: pivotal role for old-style public health measures in the novel coronavirus (2019-nCoV) outbreak. J Travel Med 2020; 27(2).

22. Perra N. Non-pharmaceutical interventions during the COVID-19 pandemic: A review. Phys Rep 2021; 913: 1–52.

23. Ahlers M, Aralis H, Tang W, Sussman JB, Fonarow GC, Ziaeian B. Non-pharmaceutical interventions and covid-19 burden in the United States: retrospective, observational cohort study. BMJ Medicine 2022; 1(1): e000030.

24. Kadri SS, Sun J, Lawandi A, et al. Association Between Caseload Surge and COVID-19 Survival in 558 U.S. Hospitals, March to August 2020. Ann Intern Med 2021; 174(9): 1240–51.

25. Luo Q, O’Connell DL, Yu XQ, et al. Cancer incidence and mortality in Australia from 2020 to 2044 and an exploratory analysis of the potential effect of treatment delays during the COVID-19 pandemic: a statistical modelling study. Lancet Public Health 2022; 7(6): e537–e48.

26. Appleby J. NHS sickness absence during the covid-19 pandemic. BMJ 2021; 372: 471.

27. Abbas M, Zhu NJ, Mookerjee S, et al. Hospital-onset COVID-19 infection surveillance systems: a systematic review. J Hosp Infect 2021; 115: 44–50.

28. Lewnard JA, Hong VX, Patel MM, Kahn R, Lipsitch M, Tartof SY. Clinical outcomes associated with SARS-CoV-2 Omicron (B.1.1.529) variant and BA.1/BA.1.1 or BA.2 subvariant infection in southern California. Nat Med 2022.

29. Nyberg T, Ferguson NM, Nash SG, et al. Comparative analysis of the risks of hospitalisation and death associated with SARS-CoV-2 omicron (B.1.1.529) and delta (B.1.617.2) variants in England: a cohort study. Lancet 2022; 399(10332): 1303–12.

30. Stratil JM, Biallas RL, Burns J, et al. Non-pharmacological measures implemented in the setting of long-term care facilities to prevent SARS-CoV-2 infections and their consequences: a rapid review. Cochrane Database Syst Rev 2021; 9: CD015085.

31. Cheung PH, Chan CP, Jin DY. Lessons learned from the fifth wave of COVID-19 in Hong Kong in early 2022. Emerg Microbes Infect 2022; 11(1): 1072–8.

32. Kwan R. How Hong Kong’s vaccination missteps led to the world’s highest covid-19 death rate. BMJ 2022; 377: o1127.

33. Yao Y, Tian J, Meng X, Kan H, Zhou L, Wang W. Progression of severity in coronavirus disease 2019 patients before treatment and a self-assessment scale to predict disease severity. BMC Infect Dis 2022; 22(1): 409.

