## Supplementary materials for "Hospital strain and Covid-19 fatality: analysis of English nationwide surveillance data"

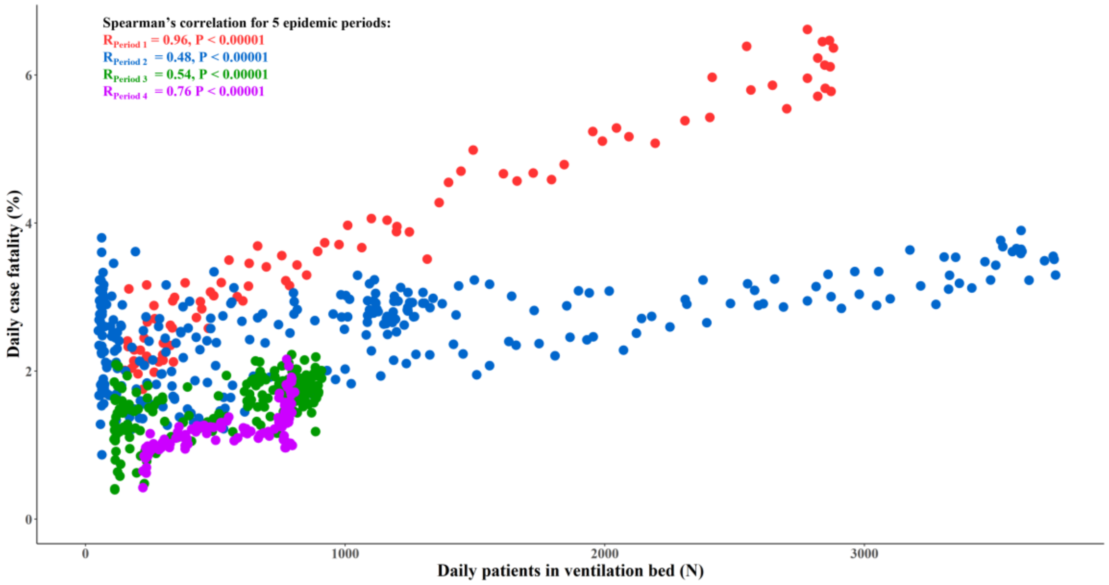


**Appendix Figure 1. The correlation of COVID-19 fatality with patients currently in ventilation beds during the 4 epidemic periods in England.**

**Appendix Table 1. Regression coefficients and relative increase in daily case fatality for each predictor in the multivariate log linear regression model in Table 3.**

| Variables | Regression coefficients (95% CI) | Relative increase (95% CI) | P-value |
|---|---|---|---|
| **Study periods** | | | |
| Period 1 | -0.005(-0.113,0.103) | 0.995(0.893,1.108) | 0.9262 |
| Period 2 | Ref (0.000) | Ref (1.000) | - |
| Period 3 | -0.180(-0.284,-0.076) | 0.835(0.753,0.927) | 0.0007 |
| Period 4 | 0.544(0.347,0.741) | 1.723(1.414,2.098) | <0.0001 |
| **Daily number of patients in hospital (per 1000 increase) in period 2** | 0.014(0.128,0.015) | 1.014(1.137,1.015) | <0.0001 |
| **Interaction term between daily number of patients in hospital (per 1000 increase) and study periods** | | | |
| Period 1 | 0.047(0.043,0.052) | - | <0.0001 |
| Period 2 | Ref (0.000) | - | - |
| Period 3 | 0.106(0.088,0.123) | - | <0.0001 |
| Period 4 | -0.019(-0.028,-0.011) | - | <0.0001 |
| **Vaccination score (per 10% increase)** | -0.051(-0.059,-0.044) | 0.95(0.943,0.957) | <0.0001 |
| **Admission rate (per 10% increase)** | -0.019(-0.040,0.002) | 0.981(0.961,1.002) | 0.0826 |
| **Percentage of death outside hospital (per 10% increase)** | -0.018(-0.045,0.009) | 0.982(0.956,1.009) | 0.1914 |

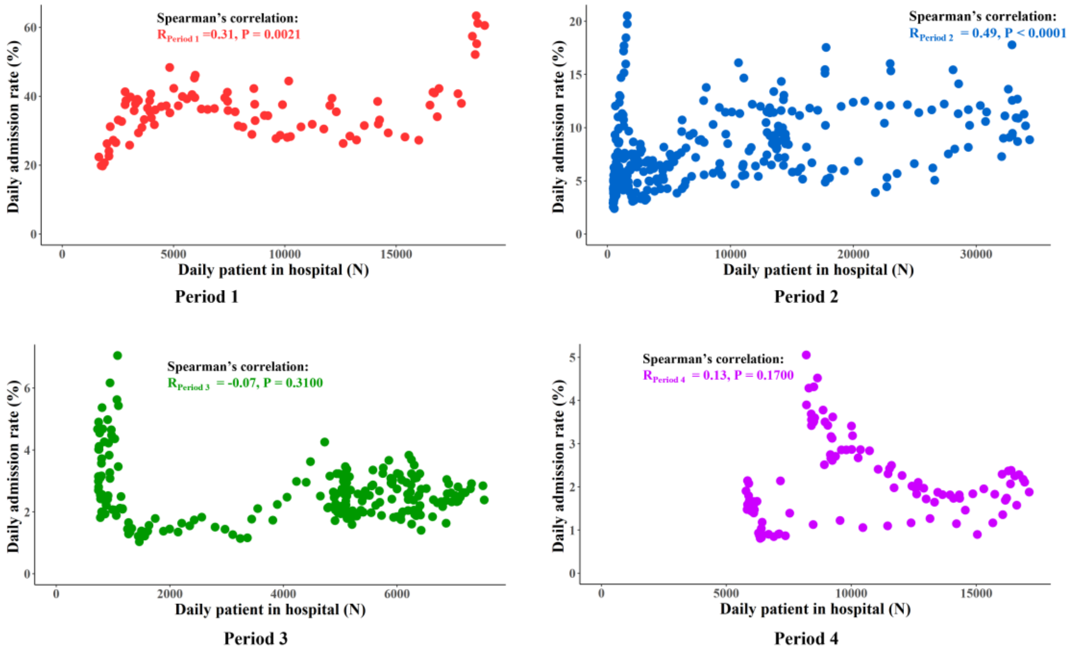


**Appendix Figure 2. The correlation of daily admission rate with patients currently in hospital during the 4 study periods.**

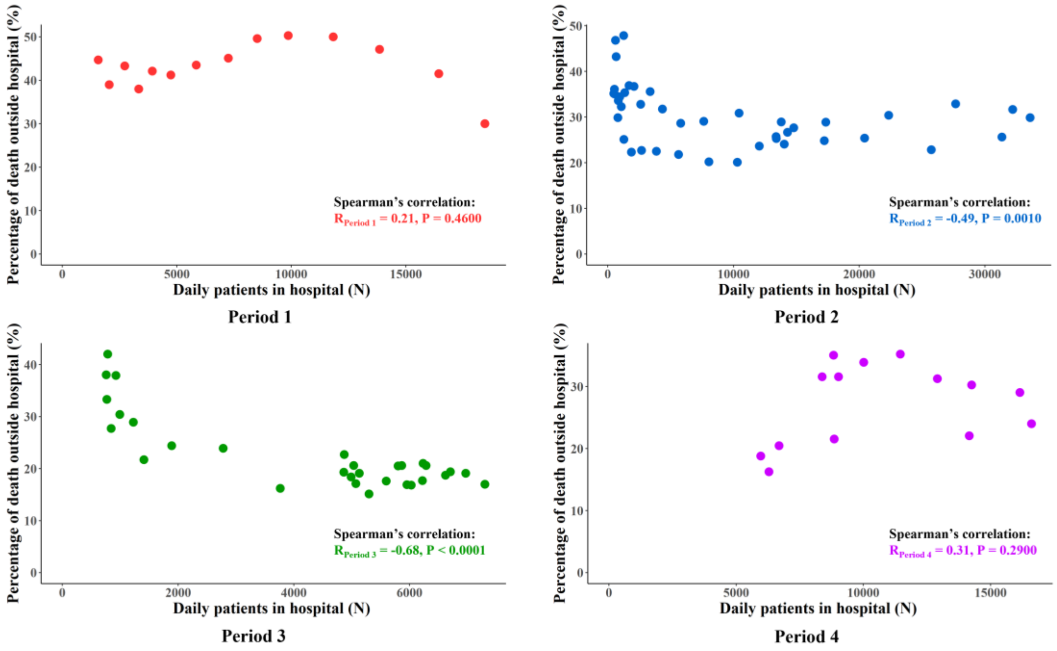


**Appendix Figure 3. The correlation of the percentage of deaths outside hospitals with daily patients currently in hospital during the 4 study periods (on a weekly basis).**
